# Spatiotemporal Modelling of Bacterial Meningitis Outbreaks using Climatic Drivers in Africa

**DOI:** 10.64898/2026.09.14.26363075

**Authors:** Molly Cliff, Mariana Perez Duque, Henrik Salje, Andre Bita Fouda, Anderson Latt, Clement Lingani, Ezra Gayawan, Caroline Trotter

## Abstract

The African meningitis belt, spanning 26 countries, experiences regular seasonal outbreaks of bacterial meningitis. Outbreaks predominantly occur during the October-April dry season, characterised by low humidity, high temperatures, and increased dust levels driven by the movement of the Harmattan winds. Whilst many studies have examined the relationship between bacterial meningitis outbreaks and climate across Africa, this has rarely been examined on a continental level. Additionally, previous continental analyses have been unable to account for spatiotemporal changes in bacterial meningitis epidemiology. We developed a district-level Bayesian spatiotemporal model to estimate monthly meningitis risk across Africa using climatic variables.

For district-months between 2003-2022 we generated a Boolean variable to identify districts affected by meningitis outbreaks, defined as an incidence of ≥10 suspected cases per 100,000 population per week. Covariates included rainfall, humidity, windspeed and direction, MenAfriVac campaign status, population density, aerosol optical depth (AOD), temperature and land cover type. We used a Bayesian Integrated Nested Laplace Approximation (INLA) model to account for spatial and temporal heterogeneity. Covariate inclusion was decided using the Watanabe Akaike Information Criterion (WAIC), the Deviance Information Criterion (DIC), as well as considering variable epidemiological relevance. Our final model included linear effects for humidity, AOD, zonal wind strength and direction, an interaction term between zonal wind direction and AOD, and a non-linear temperature effect.

Zonal wind strength had a statistically significant positive relationship with outbreaks whilst humidity had a negative association. The interaction term between zonal wind direction and AOD had a highly positive association with outbreak occurrence. When zonal wind moved westwards, higher AOD levels increased the likelihood of outbreak occurrence. Furthermore, whilst the log odds of meningitis outbreak occurrence remained low at cooler temperatures, the effect peaked between 37–38°C before beginning to plateau. The non-linear association between meningitis incidence and temperature is suggestive of an optimal temperature for bacterial transmission. Additionally, the significance of the AOD and wind direction interaction highlights the role of the Harmattan winds in exacerbating meningitis risk, rather than outbreaks solely being driven by higher dust levels. Despite these variables all having a well-established relationship with meningitis outbreaks, our study shows that such climatic associations are still present at the continental level, when accounting for spatiotemporal effects

## Introduction

Bacterial meningitis occurs when bacterial infection results in inflammation of the meninges, the layers of membrane protecting the brain and spinal cord [1]. Globally bacterial meningitis in children and young adults is predominantly caused by *Neisseria meningitidis (N. meningitidis)*, *Streptococcus pneumoniae (S. pneumoniae)* with *Haemophilus influenzae* (*Hi*) representing a significant burden in children under five [2]. Whilst these bacteria are usually commensal in humans, they can become invasive and cause disease by entering the bloodstream. This then develops into meningitis when bacteria cross the blood-brain barrier and multiply within the subarachnoid space, causing inflammation of the meninges [3]. Bacterial meningitis represents a considerable global health challenge, with an estimated 2.51 million cases having occurred in 2019. Of these, 1.28 million cases are thought to occur in children under the age of 5 [4]. Most cases occur in low- and middle-income countries where the financial burden caused by bacterial meningitis and resulting sequelae results in significant health inequity. Geographic disparity in the occurrence of bacterial meningitis is driven by regional patterns in climatic factors alongside additional socioeconomic factors, including vaccination coverage, population immunity and healthcare accessibility [5].

Globally, the highest incidence of bacterial meningitis lies within the African meningitis belt, which comprises 26 countries stretching from Senegal to Ethiopia. In this region, meningitis outbreaks are predominantly caused by *N. meningitidis,* with irregular epidemics occurring every 8 -12 years [6]. Epidemic incidence is highly seasonal with increases during the dry season (October-April), in line with the movement of the Harmattan winds, which bring Saharan dust, primarily from the Bodele Depression in Chad, west towards the Gulf of Guinea [7]. Meningitis outbreaks typically diminish with the onset of the monsoonal rains, peaking between July and September [7, 8].

Whilst many studies have examined the relationship between bacterial meningitis outbreaks and climate, few have done so at a continental level [7, 9, 10, 11, 12]. This provides the distinct advantage of capturing large-scale climatic drivers of meningitis, which could inform the location of preventative vaccination campaigns. Within our previous continental analysis, we evaluated changes in the spatial epidemiology of bacterial meningitis in the 20 years since Molesworth et al’s 2003 study [9,10]. Building on our previous analysis, our current study investigates how the relationship between climatic factors and meningitis has evolved across Africa over space and time. Alongside existing climatic relationships, the introduction of MenAfriVac, a meningococcal group A vaccine, in 2010 in West Africa is likely to have had a significant impact on the spatiotemporal heterogeneity of meningitis outbreak occurrence [13]. Additionally, some studies have found district and sub-district level clustering of meningitis cases within Africa, not accounted for in previous continental analyses [14, 15, 16]. This necessitates a more detailed examination of the spatiotemporal patterns of the outbreaks of bacterial meningitis across the African meningitis belt.

This study aims to develop a spatiotemporal model to estimate monthly meningitis outbreak risk (2003-2022) based on climate variables across the African meningitis belt. Within this study, we utilise the Integrated nested Laplace approximation (INLA) approach to account for spatial and temporal heterogeneity in outbreak occurrence. INLA has been used to undertake spatiotemporal modelling of a wide range of pathogens, including Ebola, dengue fever and Japanese encephalitis [17,18, 19]. INLA has not previously been used to model the epidemic occurrence of bacterial meningitis. Our large-scale analysis examining the monthly risk of bacterial meningitis outbreaks at a district scale can elucidate potential changes in outbreak epidemiology, aiding the management of resources in a scarce funding landscape. Surpassing previous continental-scale analyses, our model explicitly captures long-term temporal dynamics alongside spatial heterogeneity in outbreak risk.

## Materials and Methods

### Outcome data and epidemiological processing

As in Cliff et al. [9], weekly bacterial meningitis outbreak occurrence data were provided from 2003 to 2022 by the World Health Organisation Regional Office for Africa (WHO-AFRO) [20]. This data represents meningitis epidemics reported to WHO-AFRO’s enhanced meningitis surveillance system, reporting disease occurrence across sub-Saharan Africa in countries which are at high risk of meningitis outbreaks, namely Benin, Burkina Faso, Burundi, Cameroon, Central African Republic, Chad, Côte d’Ivoire, Democratic Republic of Congo, Ethiopia, Ghana, Gambia, Mali, Mauritania, Niger, Nigeria, Senegal, South Sudan, Sudan and Togo. In this analysis, we included outbreak data from all countries except the Democratic Republic of the Congo (DRC), due to its comparatively low laboratory case confirmation rates and higher levels of data duplication [9,20].

In line with updated recommendations from the World Health Organisation guidelines, in the post-MenAfriVac era, the epidemic threshold was defined as 10 suspected meningitis cases per 100,000 population per week [21]. This epidemic threshold is based on suspected cases. As such, outbreaks are therefore not identified as being specific to a particular causative pathogen. This means that whilst we assume that most outbreaks are due to meningococcal meningitis, we cannot exclude the contribution of other meningitis-causing pathogens such as pneumococcus and *Hi*.

The raw outbreak data was cleaned in Stata 18 SE, with outbreak metadata including the number of cases, deaths, attack rate per 100,000 population, ADMN2 location of outbreak, as well as week and year of outbreak. For this analysis, we assigned all outbreaks to an ADMN district within a district-level shapefile of the African continent from the Database of Global Administrative Areas (GADM) [22]. This shapefile was used to set up the primary data frame onto which the outbreak data was merged. Weekly outbreaks were temporally aggregated and assigned to their corresponding month and year of occurrence. We assumed that there was no temporal delay in outbreak reporting, as this is difficult to quantify, but acknowledge that outbreak reporting is often affected by sociopolitical factors.

In terms of spatial variance in outbreak reporting, district boundaries frequently change over time, making it more difficult to accurately assign older outbreaks to current administrative units. As with Cliff et al. [9] we used a historical ADMN2 shapefile of Africa to map bacterial meningitis epidemics to the 2022 GADM shapefile using R version 4.4.2. For outbreaks that could only be assigned to a district with the older WHO shapefile, we calculated spatial intersections between the WHO and GADM ADMN2 shapefiles and assigned outbreaks accordingly. For each month between 2003 and 2022, over all ADMN2 districts, we generated a Boolean outbreak variable to identify districts affected by outbreaks of bacterial meningitis. Districts reporting meningitis outbreaks within a given month were coded as 1, with district-months not experiencing an outbreak coded as 0.

### Environmental data

For this analysis, we examined the impact of explanatory variables, known to influence the spatiotemporal distribution of bacterial meningitis outbreaks. All environmental data used were publicly available and downloaded on a monthly level from 2003 to 2022 (S1 Table). We examined the impact of rainfall (mm), specific humidity (kg/kg), wind (m/s) (speed, meridional and zonal components), aerosol optical depth (AOD) (unitless measure), temperature (K) and land cover type on the occurrence of meningitis outbreaks [23,24,25,26,27]. Land use data from the land use harmonization2 dataset was separated into broader barren/forest/cropland categories, in line with land categories known to be associated with bacterial meningitis incidence [10,11]. Alongside the discussed climatic variables, we also examined the impact of both population density and MenAfriVac vaccination campaign occurrence on district-level meningitis outbreak risk. Vaccination campaign start dates were provided by WHO-AFRO, ADMN2 districts coded 1/0 over time if they had or had not implemented a MenAfriVac campaign.

Using the terra package [28], we masked and cropped all environmental variables to the Africa continent GADM shapefile of the African continent. We then used the package’s extract function to generate mean monthly values of all variables at the ADMN2 district level (2003-2022) across the African continent. We used both variance inflation factors (VIF) and Pearson correlation coefficients to check for collinearity between dependent variables. Within this analysis, we used a Pearson’s cut-off at 0.6 (S1 Figure) and VIF 5 as a cut-off point; no variables demonstrated any considerable collinearity.

### Space-time model set up

Within this analysis, we employed Bayesian hierarchical modelling, using the Integrated Nested Laplace Approximation (INLA) via the R-INLA package to account for spatial and temporal dependencies, which have a demonstrable influence on the heterogeneity of outbreak occurrence [29].

Our model utilised a binomial distribution, using our Boolean variable representing district months affected by an outbreak of bacterial meningitis. To determine the best-fitting model to estimate the occurrence of bacterial meningitis outbreaks across the meningitis belt, we examined several spatial and temporal random effects, assuming that outbreaks were more likely to occur in consecutive months and neighbouring districts. Our spatiotemporal selection process for the model random effects can be found in the supporting information of this paper. For both temporal and spatial random effects, performance was assessed using Deviance Information Criterion (DIC), Watanabe-Akaike information criterion (WAIC), and the logarithmic score of the Conditional Predictive Ordinate (CPO) [30] (Table 1). The WAIC and DIC are frequently used criteria for comparing and assessing model performance [30]. CPO is an internal measure of a model’s predictive ability through internal cross-validation. Low values of logCPO, WAIC and DIC are indicative of a better-fitting model. Within our spatiotemporal selection process, we opted for a Besag-York-Mollié 2 (BYM2) model to capture spatial heterogeneity and a random Walk of Order 2 (RW2) to capture the temporal random effect [31].

**Table 1:** Univariate model output of climatic and socioeconomic variable relationships with district-month level bacterial meningitis outbreaks. Continuous covariates (temperature, humidity, rainfall, meridional and zonal wind, wind speed, AOD and population density) were standardised to z scores.

| **Descriptor** | **Odds ratio**<br>**(exp($\beta$))** | **Lower**<br>**95%**<br>**Credible**<br>**interval**<br>**for log**<br>**odds**<br>**ratio** | **Higher**<br>**95%**<br>**Credible**<br>**interval**<br>**for log**<br>**odds**<br>**ratio** | **WAIC** | **Difference**<br>**to best**<br>**DIC** | **log CPO** |
| --- | --- | --- | --- | --- | --- | --- |
| Windspeed | 1.55 | 1.35 | 1.78 | 1786 | 244 | 0.001 |
| Meridional<br>wind<br>component | 0.61 | 0.53 | 0.71 | 1795 | 242 | 0.0015 |
| Zonal wind<br>component | 0.71 | 0.62 | 0.81 | 1808 | 257 | 0.0015 |
| Humidity | 1.13 | 0.82 | 1.57 | 1807 | 268 | 0.0015 |
| Temperature | 4.3 | 3.68 | 5.12 | 1625 | 0 | 0.0013 |
| MenAfriVac<br>status | 4.41 | 3.00 | 6.49 | 1828 | 242 | 0.0015 |
| Aerosol<br>optical depth<br>(AOD)- Dust | 1.99 | 1.75 | 2.27 | 1759 | 196 | 0.0014 |
| Population<br>density | 1.09 | 0.97 | 1.24 | 1809 | 272 | 0.0015 |
| Land use<br>type-<br>cropland | 1.79 | 1.00 | 3.24 | 1805 | 270 | 0.0015 |
| Rainfall | 0.28 | 0.22 | 0.35 | 1755 | 171 | 0.0014 |
| Land use<br>type- barren | 1.13 | 0.40 | 3.24 | 1811 | 273 | 0.0015 |
| Land use<br>type- forest | 0.40 | 0.18 | 0.87 | 1812 | 271 | 0.0015 |

Our final spatiotemporal random effects used to capture variation in outbreak occurrence that could not be explained by covariates follows the same structure as Salim et al., [32]. We modelled the probability of outbreak occurrence within a given district month *p_i_*_,*t*_ as:

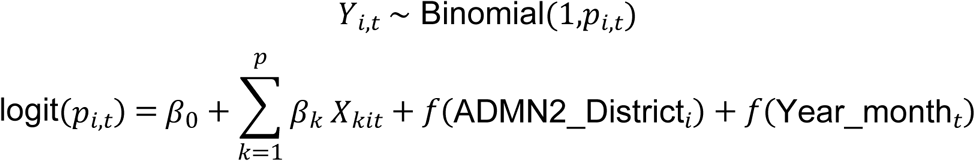

Here:

*Y_i_*_,*t*_ represents the Boolean independent variable of outbreak occurrence within a given district month

*β*_0_ is the intercept, representing baseline outbreak occurrence probability

*X_kit_*represents the climatic and socioeconomic dependent variables included in the final model.

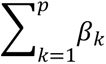 represents a sum over all the included dependent variables, with the total number of variables being represented as p. The beta values are a weighting representative. *f*(ADMN2_District*_i_*) and *f*(Year_month*_t_*) are respectively the spatial and temporal random effects.

Finally, due to the low ratio of outbreaks/non-outbreaks (affected district months = 1401/654,960) we used inverse probability weighting to adjust the contribution of each observation, allowing all data to be retained while compensating for imbalance. Whilst we also explored down-sampling of the majority (non-outbreak) class as an alternative strategy, this resulted in a significant loss of temporal and spatial information [33]. Consequently, the weighted approach was preferred, as it preserved the full spatial and temporal structure of the dataset while addressing class imbalance.

### Model selection

Initially, we conducted a univariate analysis of climatic and socioeconomic variables to check for their individual associations with meningitis outbreaks, accounting for spatiotemporal dependencies using a district-level BYM2 spatial random effects and a second-order random walk (RW2) temporal effect (Table 1). Continuous explanatory variables were standardised to z-scores in R before analysis to improve model stability and comparability of effect sizes across variables. We used changes in WAIC, DIC and logCPO alongside posterior effect estimates and 95% credible intervals from the univariate analysis to decide on variable inclusion within the full model. Within our univariate analysis we used INLA’s default priors for both the fixed and random effects due to the exploratory nature of this analysis in variable selection. A covariate was considered significantly associated with the occurrence of bacterial meningitis outbreaks if its 95% credible interval for the log odds ratio did not cross zero. Within the univariate analysis, temperature, district-level MenAfriVac campaign, windspeed and aerosol optical depth (AOD) all had a statistically significant positive association with meningitis outbreaks. Conversely, zonal and meridional wind components, forested areas and rainfall all had a statistically significant negative association with outbreak occurrence.

The final model was developed using a forward stepwise strategy with candidate variables added sequentially and retained based on improvements in WAIC and DIC as well their epidemiological relevance from relevant literature. Temperature was used as our baseline explanatory variable, as it led to the largest reduction in both WAIC and DIC within our univariate analysis. Zonal wind was subsequently included as a linear effect due to the significant reduction it caused in both DIC and WAIC. AOD was also incorporated due to its epidemiological relevance and relationship with zonal wind in driving the movement of the Harmattan winds carrying dust across West Africa. Although humidity did not show a statistically significant association in the univariate model, its inclusion was considered due to its predictive capability in previous papers [9,10,34] .

Potential non-linear relationships were evaluated using RW2 effects for included explanatory variables. Including a RW2 effect for temperature led to the largest reduction in both WAIC and DIC and was included within the final model. We examined potential interactions between zonal wind and AOD to capture the movement of the Harmattan winds across West Africa. As part of this, we extracted the absolute values of the zonal wind speed variable to represent directional wind strength and translated the zonal wind variable into a binary directional indicator to represent easterly and westerly wind flow. The interaction between dust and binary wind direction led to a notable reduction in both WAIC and DIC and was included in the final model. In contrast, the interaction between dust and wind strength showed limited improvement and was removed from the final model to aid model parsimony. Within our final multivariable model, we used weakly informative priors Normal (mean = 0, precision = 1), assuming that covariates were likely to have a limited effect on outbreak occurrence given their rarity. For all random effects we used non-informative penalised complexity (PC) priors, which penalise departure from a simple baseline model, unless there is supporting evidence. For both the RW2 temporal and temperature effects we set the precision hyperparameters at (0.5, 0.01) . For the BYM2 spatial random effect we set the precision hyperparameter at (1,0.01) and the mixing parameter at (0.5, 0.6), aiming to balance structured and unstructured spatial variations. Within our SI we carried out an additional analysis, demonstrating model sensitivity to choices in priors, examining the impact on both fixed effects, WAIC and DIC.

### Cross validation and model evaluation

We carried out stratified k fold cross validation of our model over 5 separate folds. Due to the significant variation in outbreak incidence over both space and time we ensured that outbreaks were equally balanced across different folds. We avoided data leakage by making sure that outbreaks in sequential months within the same ADMN district were contained within the same fold. Each of the five folds was held out in turn and the model was trained on the remaining four folds. The model trained on the four test folds was then asked to predict onto the test fold. Predicted values across the five folds were joined into an out of fold dataset used for model evaluation.

Model performance was evaluated using the area under the receiver operator curve (AUC ROC) as well as the Brier score and log loss, which are both measures of how close predicted probabilities are to actual outcomes, with log loss having strong penalization for confident but false predictions. Both the Brier score and log loss are strictly proper scoring rules where lower values are indicative of a better fitting model. For both tests a perfect score would be 0 with the worst score being 1. Alongside this, we evaluated model accuracy using precision and recall across different cut off thresholds.

The repository for this code can be found on GitHub via the following link https://github.com/molly-cliff/Continental_African_Bayesian_Analysis

## Results

### Geographic and Temporal Distribution of Meningitis Outbreaks

When grouping together districts affected by bacterial meningitis outbreaks across concurrent months and neighbouring locations, we found 330 separate outbreak events across 19 countries. The Central and Western part of the Sahelian region suffered the highest burden of meningitis outbreaks (Figure 1a). Burkina Faso, Chad, Niger and Nigeria reported that >50% of their ADMN2 districts were affected by an outbreak, demonstrative of localised spatial clustering. Across the belt, several districts were repeatedly affected by distinct outbreak events, highlighting regional vulnerability to disease. Tanguiéta in Benin reported 12 outbreaks over 8 years, with districts across Burkina Faso also reporting a high number of outbreaks.

**Figure 1:**
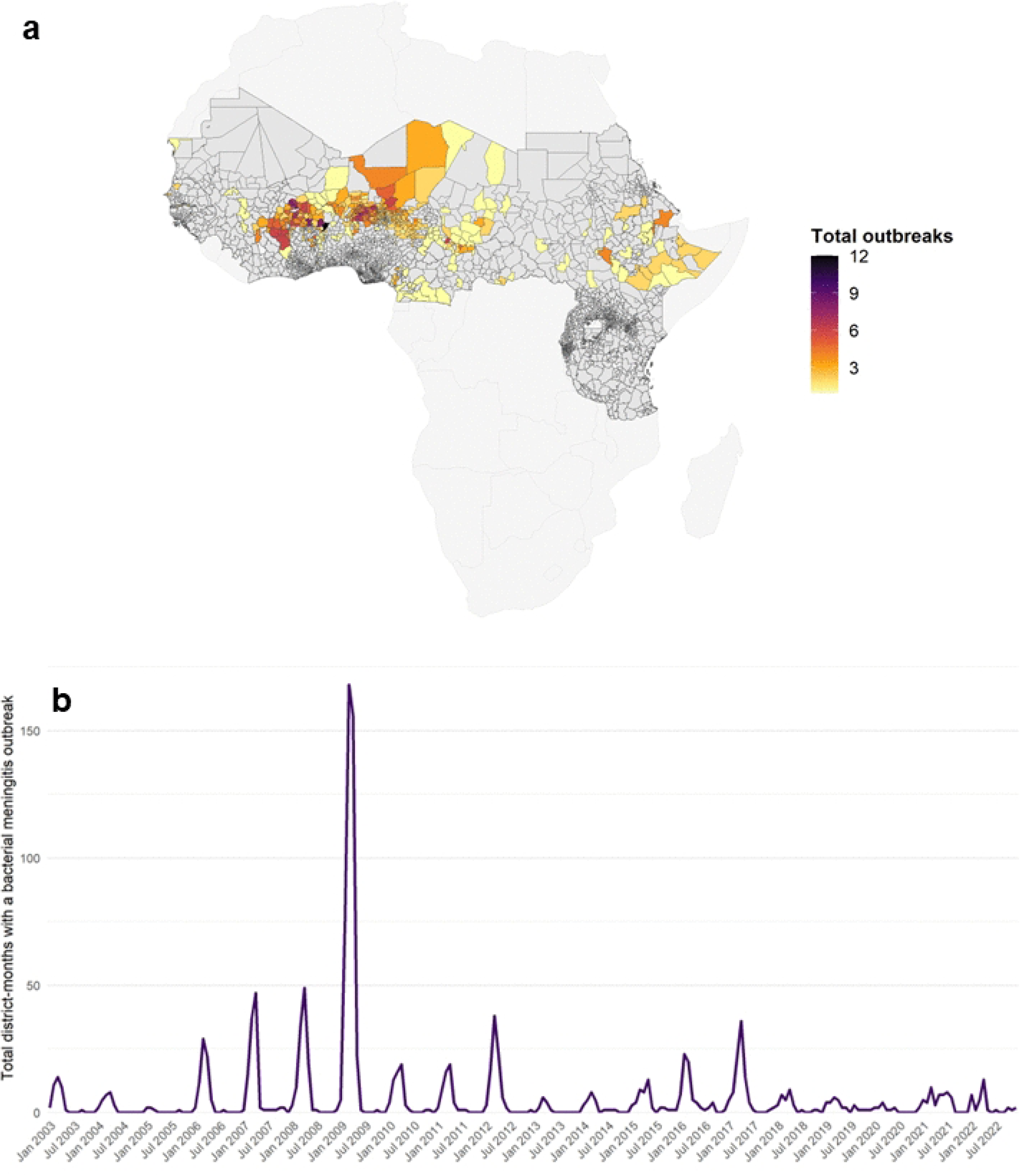
a: Map of number of bacterial meningitis epidemics across reporting countries (2003–2022) b: Time series graph showing the total number of district-months affected by a bacterial meningitis outbreak over time.

Across the 20-year surveillance period the largest outbreak event occurred in West Africa between 2008-2009, affecting Burkina Faso, Mali, Niger and Nigeria (Figure 1b). Within this outbreak there were 165 ADMN2 districts affected within a single month, three times larger than any other single outbreak event. Since the introduction of MenAfriVac in 2010 there has been a notable reduction in outbreak magnitude and frequency, with epidemics post 2018 rarely exceeding 10-15 district-months. Across the study time frame, epidemics typically occurred within the October to May dry season in recurrent months, with smaller intermittent outbreaks occurring outside of this.

### Bayesian model linear effects

Of all the models evaluated, model 12 (Table 2) had the lowest WAIC and DIC, suggesting better goodness of fit. As with our previous models, this iteration included random effects for both space (BYM2) and time (RW2), but additionally modelled temperature through a non-linear RW2 effect. Our final model included a random effect for temperature, alongside a linear term for specific humidity and an interaction term between zonal wind direction and AOD. Alongside this the model included district-level spatial (BYM2) and monthly temporal random effects (RW2).

**Table 2:**
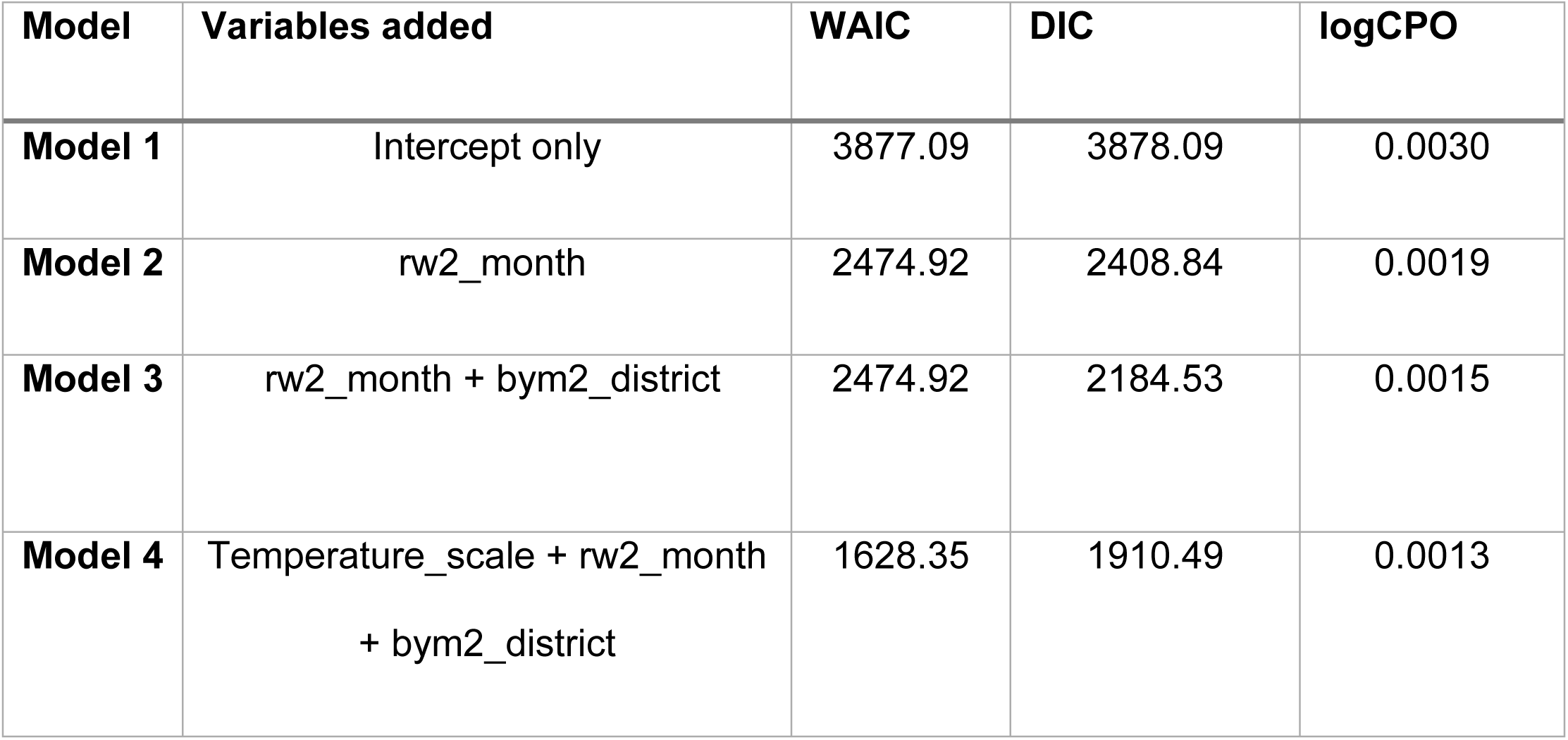
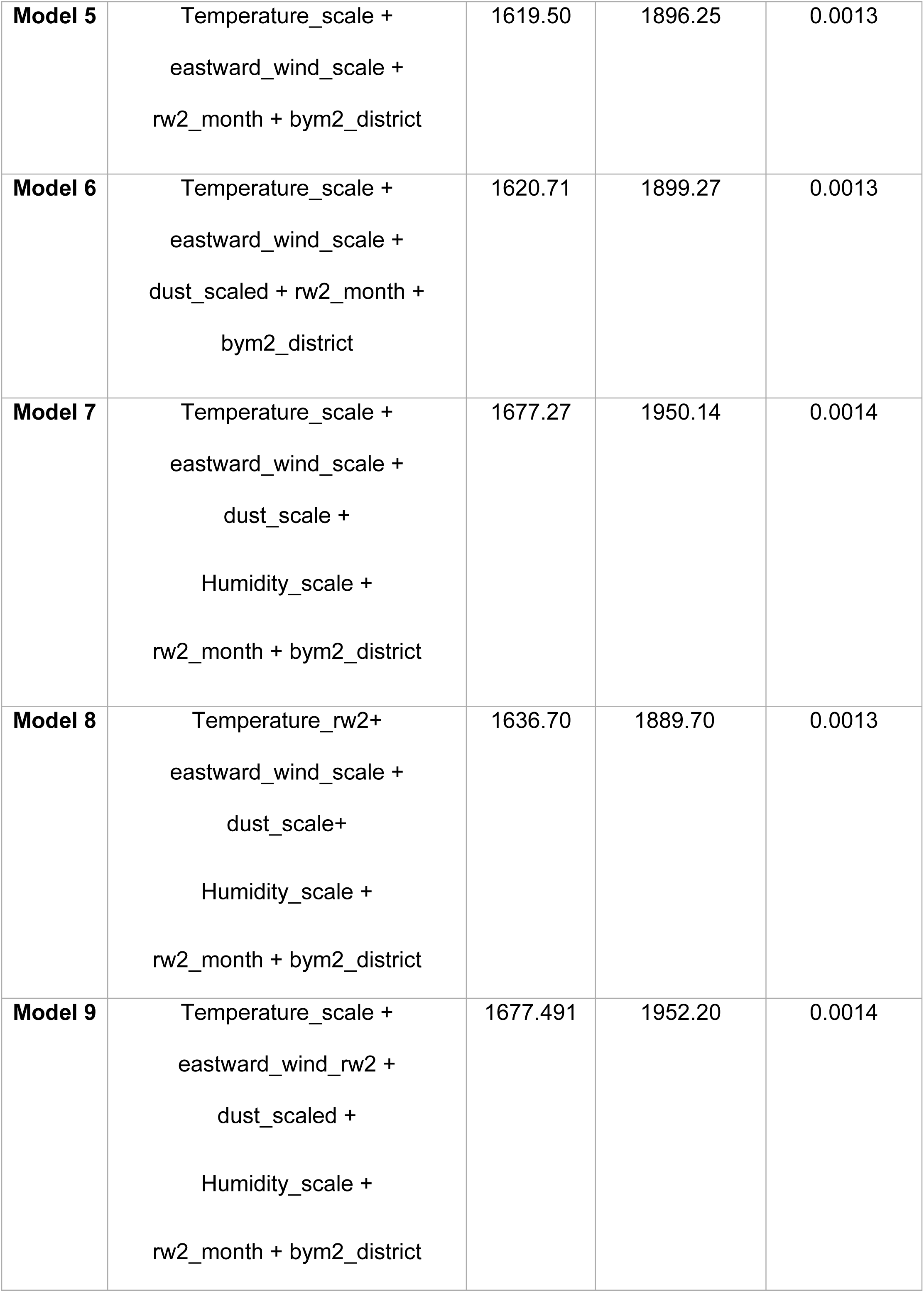
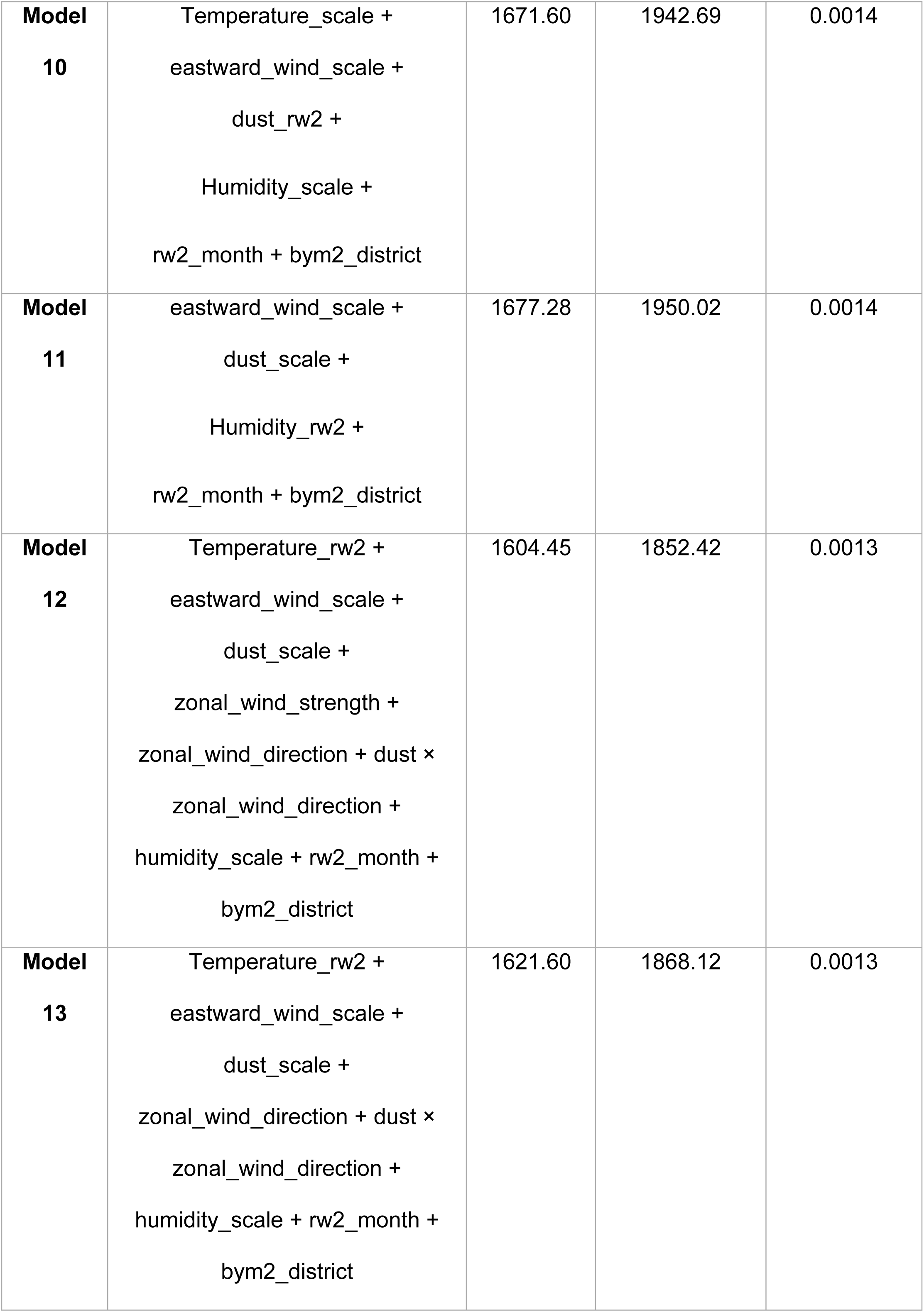
Table showing formulae of tested INLA models alongside their WAIC, DIC and logCPO.

| Model | Variables added | WAIC | DIC | logCPO |
| --- | --- | --- | --- | --- |
| Model 1 | Intercept only | 3877.09 | 3878.09 | 0.0030 |
| Model 2 | rw2_month | 2474.92 | 2408.84 | 0.0019 |
| Model 3 | rw2_month + bym2_district | 2474.92 | 2184.53 | 0.0015 |
| Model 4 | Temperature_scale + rw2_month<br>+ bym2_district | 1628.35 | 1910.49 | 0.0013 |
| **Model 5** | Temperature_scale +<br>eastward_wind_scale +<br>rw2_month + bym2_district | 1619.50 | 1896.25 | 0.0013 |
| **Model 6** | Temperature_scale +<br>eastward_wind_scale +<br>dust_scaled + rw2_month +<br>bym2_district | 1620.71 | 1899.27 | 0.0013 |
| **Model 7** | Temperature_scale +<br>eastward_wind_scale +<br>dust_scale +<br>Humidity_scale +<br>rw2_month + bym2_district | 1677.27 | 1950.14 | 0.0014 |
| **Model 8** | Temperature_rw2+<br>eastward_wind_scale +<br>dust_scale+<br>Humidity_scale +<br>rw2_month + bym2_district | 1636.70 | 1889.70 | 0.0013 |
| **Model 9** | Temperature_scale +<br>eastward_wind_rw2 +<br>dust_scaled +<br>Humidity_scale +<br>rw2_month + bym2_district | 1677.491 | 1952.20 | 0.0014 |
| **Model**<br>**10** | Temperature_scale +<br>eastward_wind_scale +<br>dust_rw2 +<br>Humidity_scale +<br>rw2_month + bym2_district | 1671.60 | 1942.69 | 0.0014 |
| **Model**<br>**11** | eastward_wind_scale +<br>dust_scale +<br>Humidity_rw2 +<br>rw2_month + bym2_district | 1677.28 | 1950.02 | 0.0014 |
| **Model**<br>**12** | Temperature_rw2 +<br>eastward_wind_scale +<br>dust_scale +<br>zonal_wind_strength +<br>zonal_wind_direction + dust ×<br>zonal_wind_direction +<br>humidity_scale + rw2_month +<br>bym2_district | 1604.45 | 1852.42 | 0.0013 |
| **Model**<br>**13** | Temperature_rw2 +<br>eastward_wind_scale +<br>dust_scale +<br>zonal_wind_direction + dust ×<br>zonal_wind_direction +<br>humidity_scale + rw2_month +<br>bym2_district | 1621.60 | 1868.12 | 0.0013 |

When examining the linear fixed effects of the final model (Table 3) the strongly negative intercept reflects the rarity of outbreaks despite inverse probability weighting. However, the results highlight both the positive association that zonal wind strength has with incidence of meningitis outbreaks (posterior mean ≈ 0.24, 95% credible interval 0.11 to 0.37), alongside the clear negative association of humidity (posterior mean ≈ -0.73, CrI -0.91 to -0.54). Although both AOD (posterior mean ≈ -0.19 , 95% credible interval -0.4 to 0.02) and binary wind direction (posterior mean ≈ -0.074, 95% credible interval -0.38 to 0.24) individually do not have a statistically significant effect on epidemic incidence, when considering both effects together, the interaction has a strong positive association with bacterial meningitis outbreaks (posterior mean ≈ 0.903, 95% credible interval 0.65 to 1.15). This highlights the changing effect of dust levels on meningitis risk according to wind direction. Our results indicate that the risk of outbreak occurrence increases with higher dust levels when wind moves east to west, in line with the movement of the Harmattan winds.

**Table 3:** Fixed effect posterior estimates of final model of association of environmental factors with bacterial meningitis outbreak risk across the meningitis belt. Here continuous covariates (temperature, humidity and AOD), were standardised to z scores. Absolute values of zonal wind speed were extracted to represent directional wind strength with the zonal wind variable also translated into a binary directional indicator to represent easterly and westerly wind flow.

| Fixed Effect | Mean | Standard deviation | 2.5% Quantile | 50% Quantile | 97.5% Quantile |
| --- | --- | --- | --- | --- | --- |
| (Intercept) | -1.60 | 0.29 | -2.19 | -1.60 | -1.04 |
| aod_scale | -0.19 | 0.11 | -0.40 | -0.19 | 0.02 |
| zonal_wind_strength | 0.24 | 0.07 | 0.11 | 0.24 | 0.37 |
| Humidity_scale | -0.73 | 0.09 | -0.91 | -0.73 | -0.54 |
| zonal_wind_direction | -0.074 | 0.16 | -0.38 | -0.07 | 0.24 |
| aod_scale:zonal_wind_direction | 0.90 | 0.18 | 0.65 | 0.90 | 1.15 |

### Non-linear effects and interaction

Figure 2a highlights the non-linear effect for temperature, demonstrating how the log odds of outbreak occurrence changes as temperatures rise. For interpretability whilst the original temperature data was in Kelvin (K) we converted this to Celsius (°C). Although the log odds of outbreak occurrence remain relatively low at cooler temperatures, the no effect line is crossed at 32°C. At this point, the log odds of outbreak incidence increase until 37°C, whereby the risk begins to plateau. This is supported by Figure 2b, demonstrating the occurrence of bacterial meningitis outbreaks across a range of temperatures. Notably, most outbreaks occur between 35-38°C, with limited observations at lower temperatures. Figure 3 shows the changing log odds of outbreak occurrence as described by the interaction term between zonal wind direction and AOD. When zonal wind flows east to west, the log odds of outbreak occurrence increase with higher AOD levels. Conversely, when wind flows west to east, the log odds of outbreak occurrence decrease as AOD levels rise.

**Figure 2.**
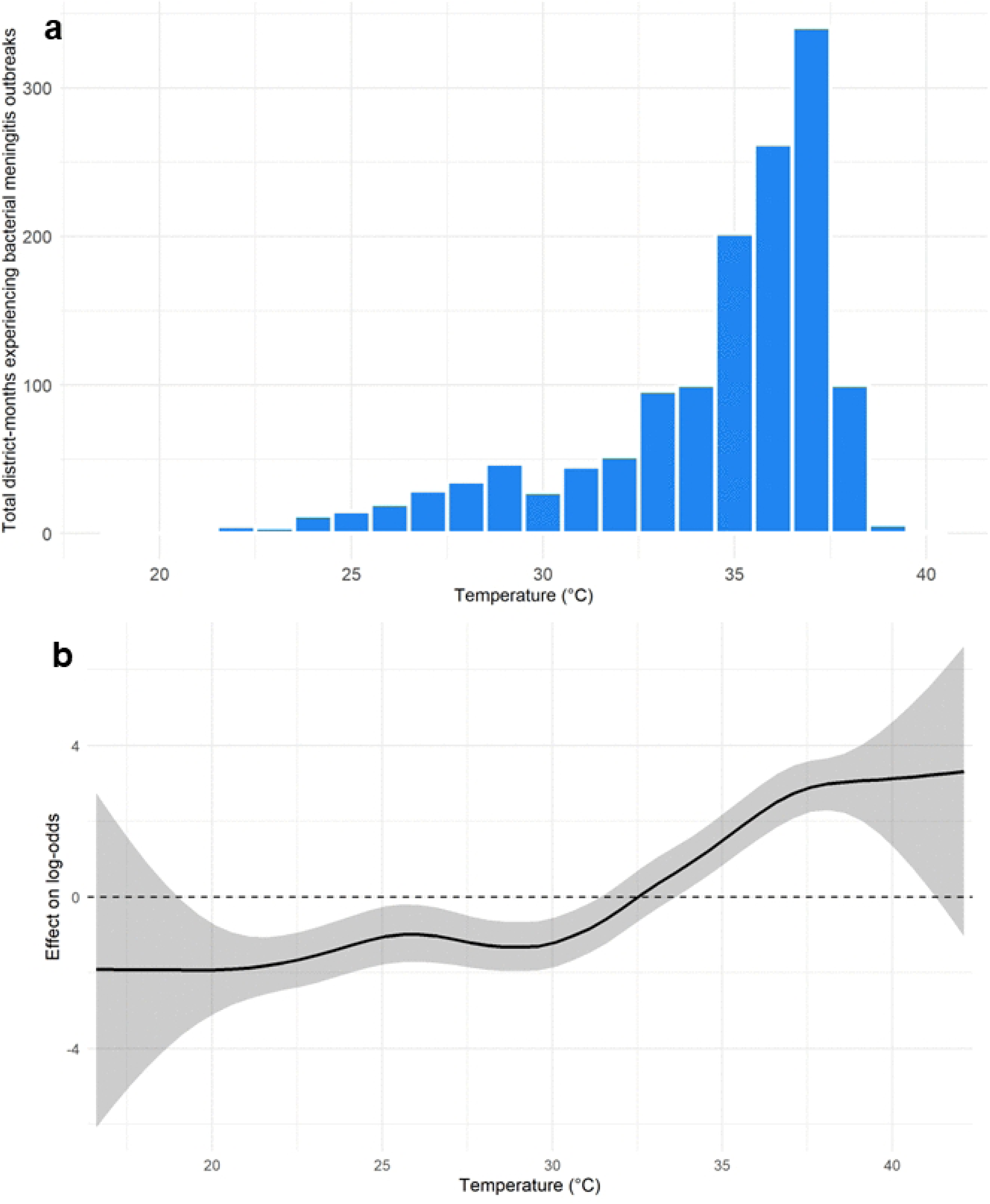
a: RW2 extracted effect of temperature showing how change affects log odds of outbreak occurrence. Here the grey ribbon represents the 95% credible interval. b: Histogram showing the total number of district-months affected by a bacterial meningitis outbreak by temperature.

**Figure 3:**
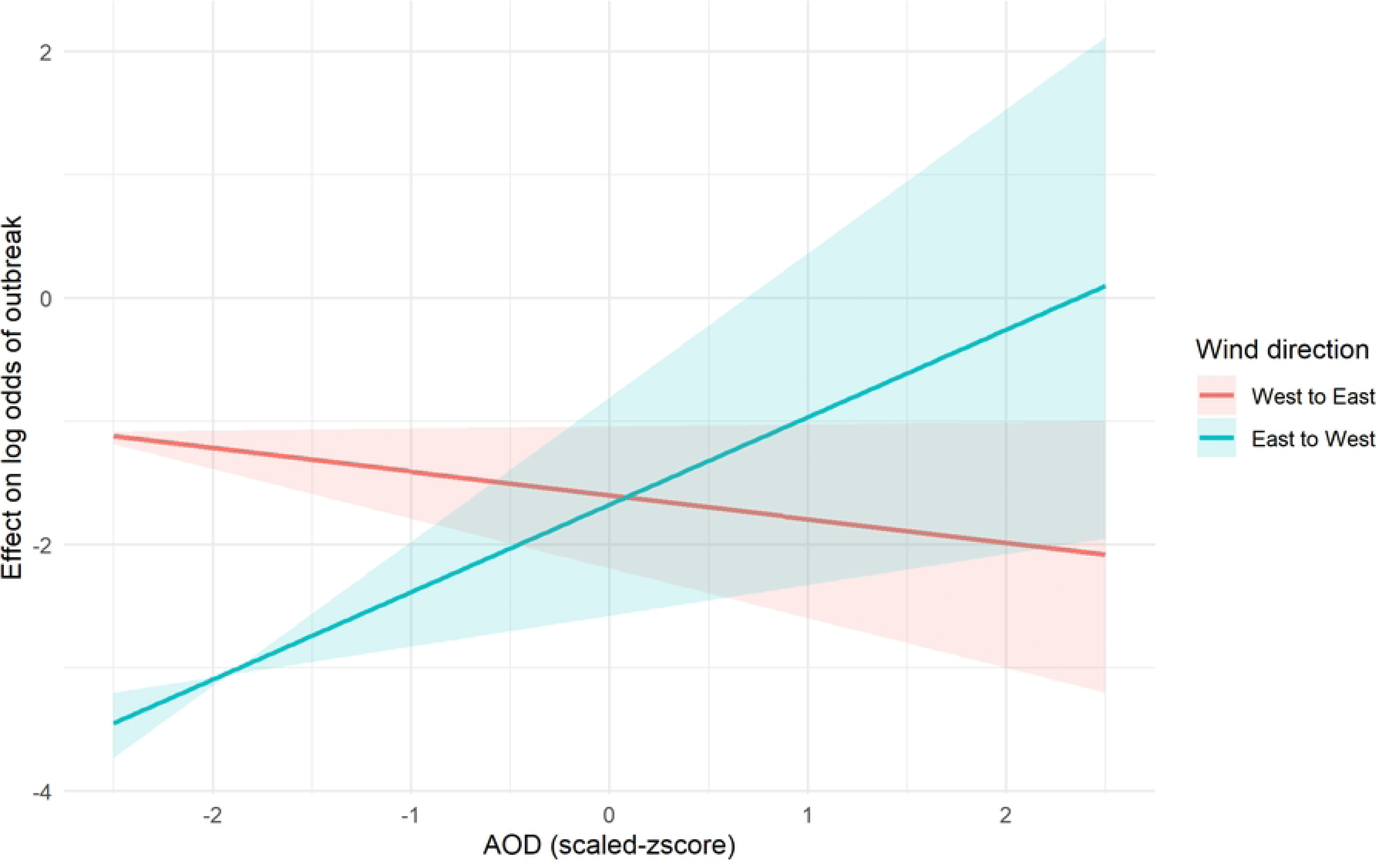
Extracted linear effect of interaction between wind direction and AOD. This graph shows how under different wind directions the effect of increasing dust changes the log odds of outbreak occurrence.

Figure 4 shows the model’s RW2 temporal effect on meningitis outbreaks on a long-term and monthly scale. There are strong fluctuations in the temporal random effect, highlighting both increased outbreak odds, following the 2009 West Africa outbreak, as well as periods of decreased risk in 2005 and 2013. When this is averaged to a monthly scale, the model highlights the increased risk of outbreaks in March and April across the belt towards the tail end of the dry season. This then decreases to reach the lowest level of outbreak risk between August and September. The low log odds overall highlight both the rarity and stochasticity of bacterial meningitis outbreaks. The model also demonstrated some localised clustering of districts experiencing meningitis outbreaks in both East and West Africa from the spatial random effect (S3 Figure). The removal of spatial and temporal random effects from the model worsened model performance in terms of both WAIC and DIC. As such both random effects were included within the final climatic model.

**Figure 4:**
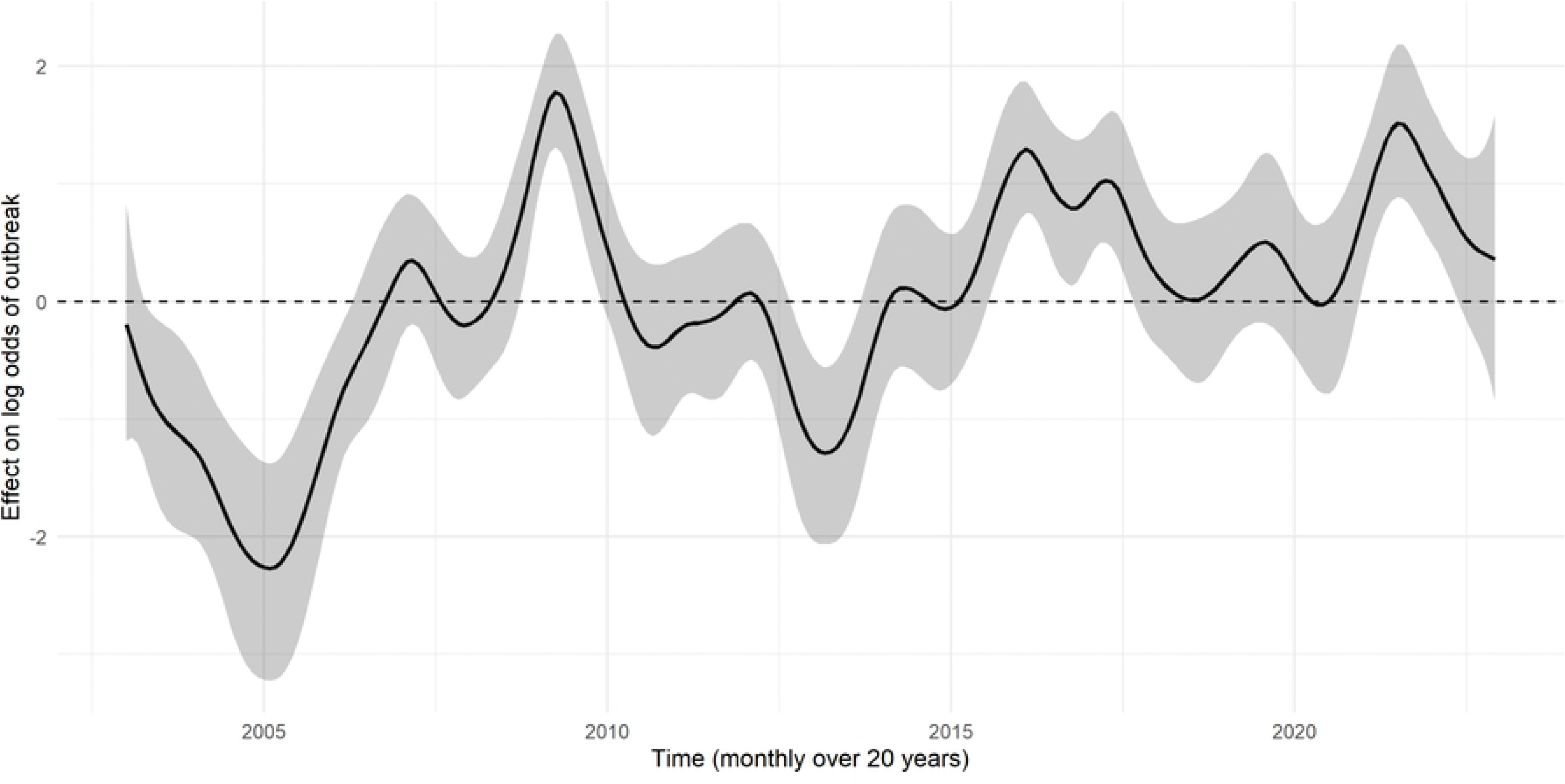
RW2 temporal effect on log odds of meningitis outbreak occurrence. Here, the grey ribbon represents the 95% credible interval.

### Model cross validation

Cross validation highlighted the model’s ability to distinguish well between district months affected by an outbreak and those not affected. The receiver operator curve was used to evaluate model performance in reference to its ability to distinguish between true positives and false positives across all possible classification thresholds. We evaluated the area under the curve (AUC) across all five folds as they were left out in turn (Figure 5). The average area under the curve (AUC) was 0.905 (0.88-0.92), demonstrating that the epidemic risk assigned was higher for district-months with epidemics than for those without in 90.5% of ADMN2 districts. When evaluating the model on how close predicted probabilities were to actual outcomes both the Brier scores (0.091-0.11) and log loss (0.31-0.36) remained relatively consistent across all five cross validation folds (Table 4).

**Figure 5:**
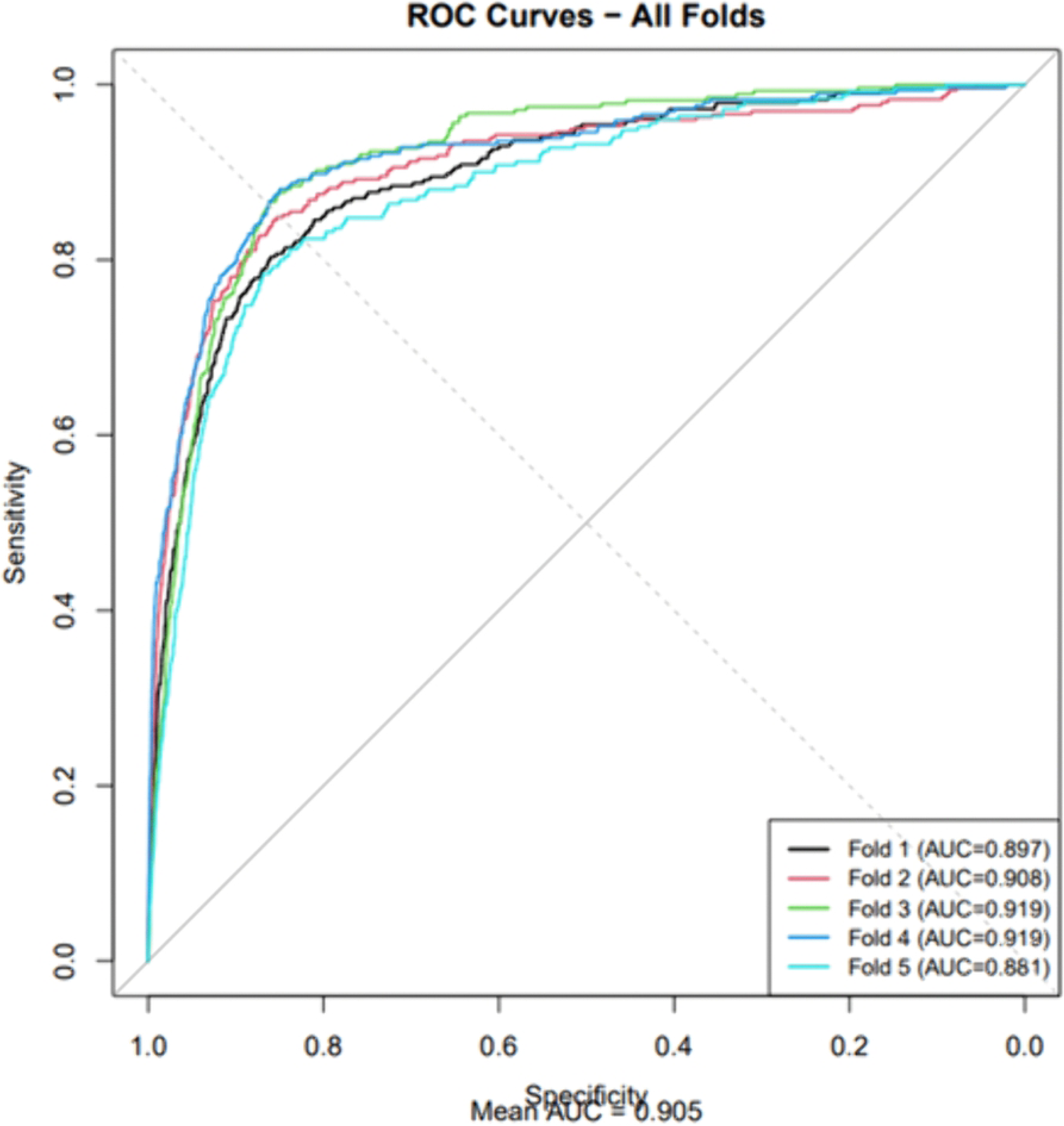
Receiver Operating Characteristic Curve for cross validation of INLA climatic model.

**Table 4:** Brier score and logloss results for cross validation of INLA climatic model.

| K fold | Brier score | Logloss |
| --- | --- | --- |
| 1 | 0.091 | 0.31 |
| 2 | 0.097 | 0.33 |
| 3 | 0.10 | 0.34 |
| 4 | 0.098 | 0.32 |
| 5 | 0.11 | 0.36 |

Table 5 highlights the average precision and recall of the climatic model on the five cross-validation folds for varying probability thresholds corresponding to the top predicted fractions of district months. With the underlying dataset both being very large highly imbalanced (affected district months = 0.214%), model precision will always be low. This means that even if model sensitivity and specificity is high there will consistently be a significant number of false positives. Across the five cross validation folds the model shows a good ability identify outbreak affected district-months, with the top 0.01% of predicted responses encompassing 24% of all outbreaks. The top 0.1% of predicted responses correspond to 75% of all outbreaks, highlighting the model’s ability to discriminate well between outbreak and non-outbreak events. This necessitates a relatively low probability cut off threshold of 0.18, consistent with the rarity of outbreaks, leading to relatively low probability estimates.

**Table 5:** Climatic INLA model precision and recall results on concatenated cross validation dataset at different cut off thresholds

| Top percentage of predicted responses (%) | Probability cut off threshold | Precision | Recall |
| --- | --- | --- | --- |
| 0.01 | 0.72 | 0.051 | 0.24 |
| 0.05 | 0.33 | 0.025 | 0.58 |
| 0.1 | 0.18 | 0.016 | 0.75 |

### Risk mapping

Finally, we present the monthly risk maps (S4 Figure-available via FigShare repository doi: 10.6084/m9.figshare.29769407) from our INLA model, demonstrating the strong seasonality in outbreak risk across meningitis belt countries. The model’s monthly outbreak risk demonstrated a high level of periodicity across our study period, with outbreak risk concentrated in the Central Sahelian predominately within Niger, Burkina Faso and Chad. Alongside this, districts in Ethiopia also had a demonstrable risk of bacterial meningitis outbreaks. Within our model, the highest risk of outbreak occurred in 2009 associated with the West Africa outbreak. Other periods of increased risk are March-April in 2017, 2019 and 2021.

## Discussion

In this study, we created a Bayesian spatiotemporal model, examining the key climatic drivers of bacterial meningitis across 19 countries in the African meningitis belt, providing large-scale validation of these relationships. Our analysis identified increasing temperature, decreasing humidity and the interaction between atmospheric dust and zonal wind direction as significantly associated with outbreak occurrence. Despite these variables all having a well-established relationship with meningitis outbreaks, our study shows that such climatic associations are still present at the continental level, when accounting for spatiotemporal effects.

While aerosol optical depth and binary zonal wind direction do not have a statistically significant effect on epidemic incidence, the interaction has a strong positive association with bacterial meningitis outbreaks. Our model suggests that across the meningitis belt, increased dust levels alone do not drive the risk of bacterial meningitis outbreaks, but this occurs in combination with the directional movement of zonal winds. When zonal wind moves east to west, the log odds of outbreak occurrence increase as dust levels rise, in line with the movement of the Harmattan winds. Such findings support the mechanistic hypothesis that atmospheric dust carried by the Harmattan winds can increase bacterial meningitis outbreak risk through increasing bacterial transmission and invasion [7, 8, 12,35, 36,37,38,39]. The limited statistical significance of both variables suggests that the mechanistic transport of atmospheric dust is more important than dust concentration or wind strength alone.

We also used a random walk 2 effect to capture how long-term variations in temperature can impact the risk of bacterial meningitis outbreak occurrence. This reinforced existing knowledge regarding outbreak association with temperature, with the log odds of outbreak occurrence remaining lower at cooler temperatures and crossing the no effect line at 32°C. The log odds of outbreak incidence increase until 37-38°C, whereby risk begins to plateau. These results are noticeably higher than the weekly mean temperature threshold of 30 °C suggested by Dione et al. [39], but are supported by observational data. Whilst some papers have found a statistically significant link between increasing temperatures and meningococcal meningitis incidence [40,41,42,43,44] the plateauing of risk at a certain threshold supports the possibility of an optimal temperature for bacterial meningitis transmission. As in Dione et al’s analysis [39], an optimal temperature threshold could be used to support existing early warning systems. The existence of an optimal temperature for bacterial meningitis transmission suggests that climate change may not result in a linear increase in bacterial meningitis incidence but could alter the geographical distribution and seasonality of outbreak risk.

However, our study has some limitations, with several epidemiological and socioeconomic variables associated with disease outbreak not being included within our model, including population migration and immunity levels. It is unlikely that these socioeconomic variables would be available on a continental level at an appropriate resolution. Data completeness also serves as an additional limitation. While the enhanced meningitis surveillance network was established in 2003, an electronic application was only introduced in 2005, with data remaining incomplete across several Sahelian countries, including Chad and Nigeria [20].

This limitation is reflected in the low log-odds of outbreak occurrence in 2005. Our model found a limited association between the occurrence of bacterial meningitis epidemics and district-level vaccination with MenAfriVac, reflecting the decreasing incidence of serogroup A meningococcal disease, coinciding with outbreaks caused by serogroups W and C [45,46]

With Gavi, the Vaccine Alliance funding having fallen short of its target following the withdrawal of US support, this necessitates the reprioritisation of outbreak resources at the continental level [47]. With the introduction of Men5CV in Niger and northern Nigeria [48], our model could be used to identify areas with persistent outbreak risk, as potential candidates for vaccination campaigns. The statistically supported non-linear temperature threshold within our study could be used to inform warning thresholds for epidemic occurrence, in line with studies from Dione et al [39] and Pandya et al [34]. Finally, due to the stochasticity in the occurrence of bacterial meningitis outbreaks, the monthly baseline estimates of district-level outbreak risk within our model could support the projection of future outbreak risk. We aim to use our INLA model to estimate the future climatic risk of meningitis outbreaks across Africa using global climate models across several shared socioeconomic pathways.

## Data Availability

Code supporting this paper is available at https://github.com/molly-cliff/Continental_African_Bayesian_Analysis, with Figure S4 (animated risk map of outbreak risk) available on FigShare via https://doi.org/10.6084/m9.figshare.29769407 Climatic data All environmental data used within this analysis is publicly available, with the data sources of each variable being detailed within the supporting information of this paper. The specific humidity, wind speed, zonal wind flow, and meridional wind flow data used are available from the European Centre for Medium-Range Weather Forecasts (ECMWF) ERA5 reanalysis on the Copernicus Climate Data Store via https://doi.org/10.24381/cds.f17050d7. The rainfall data are available from Climate Hazards Group InfraRed Precipitation with Station data (CHIRPS) via https://www.chc.ucsb.edu/data/chirps. The aerosol optical depth (dust) data are available from the National Aeronautics and Space Administration MODIS via http://dx.doi.org/10.5067/MODIS/MOD04_L2.061. Temperature data used is from NASA AIRS/Aqua L3 Monthly Standard Physical Retrieval and can be found at https://airs.jpl.nasa.gov/data/get-data/standard-data/. Land-use type data were obtained from LUH2, available at https://luh.umd.edu/. Population density data were obtained from SEDAC Gridded Population of the World Version 4, available at https://sedac.ciesin.columbia.edu/data/set/gpw-v4-population-density-rev11/data-download Epidemiological data Based on WHO-AFRO data sharing guidelines, the full epidemiological dataset underlying the findings in this manuscript cannot be shared due to the need to protect the confidentiality of district-level outbreak surveillance and MenAfriVac campaign data. These data were provided by co-authors André Bita, Anderson Latt, and Clément Lingani and are not publicly available in the format used in this publication. To access this data an application would need to be made via Clément Lingani who can be contacted at. A range of publicly available meningitis surveillance data are available via the Power BI WHO meningitis data dashboard- https://app.powerbi.com/view?r=eyJrIjoiMzExZTUzZDAtMmQ5Ni00YjkxLWFmZmItODljOWZhOTMyM2Y3IiwidCI6ImY2MTBjMGI3LWJkMjQtNGIzOS04MTBiLTNkYzI4MGFmYjU5MCIsImMiOjh9

## Acknowledgements

This work was carried out as part of the Vaccine Impact Modelling Consortium (www.vaccineimpact.org), but the views expressed are those of the authors and not necessarily those of the Consortium or its funders. The funders were given the opportunity to review this paper prior to publication, but the final decision on the content of the publication was taken by the authors. This work was primarily supported by the Wellcome Trust via the Vaccine Impact Modelling Consortium (Grant 226727_Z_22_Z), with additional support from the Gates Foundation (Grant INV-034281), previously (OPP1157270/ INV-009125), and Gavi, the Vaccine Alliance. The conclusions and opinions expressed in this work are those of the author(s) alone and shall not be attributed to the Foundation. Under the grant conditions of the Foundation, a Creative Commons Attribution 4.0 License has already been assigned to the Author Accepted Manuscript version that might arise from this submission. Please note works submitted as a preprint have not undergone a peer review process. We also acknowledge funding provided by the Jameel Institute (supported by a philanthropic donation from Community Jameel) and from the MRC Centre for Global Infectious Disease Analysis (reference MR/X020258/1), funded by the UK Medical Research Council (MRC). The latter UK-funded award is carried out in the frame of the Global Health EDCTP3 Joint Undertaking. The funders had no role in study design, data collection and analysis, decision to publish, or preparation of the manuscript.

## Supporting information captions

S1 Table: Data sources and resolution for all variables included within Bayesian INLA analysis

S1 Figure: Data sources and resolution for all variables included within Bayesian INLA analysis

S1 Text: Temporal effects exploration

S2 Table: Comparison of Bayesian model fit across different temporal model set-ups using the DIC, WAIC and logCPO criteria.

S2 Text: Spatial effects exploration

S3 Table: Comparison of Bayesian model fit across different spatial model set-ups using the DIC, WAIC and logCPO criteria.

S3 Text: Priors’ sensitivity analysis

S4 Table: Prior specification across tested models within sensitivity analysis

S2 Figure: Posterior means of fixed effects of final model variables across different prior specifications

S5 Table: Table showing WAIC and DIC of models with varying prior specifications

S3 Figure: Data sources and resolution for all variables included within Bayesian INLA analysis

S4 Figure: Monthly outbreak risk mapping across the meningitis belt

